# The effect of obesity-related traits on COVID-19 severe respiratory symptoms is mediated by socioeconomic status: a multivariable Mendelian randomization study

**DOI:** 10.1101/2021.06.08.21258587

**Authors:** Brenda Cabrera-Mendoza, Frank R Wendt, Gita A Pathak, Flavio De Angelis, Antonella De Lillo, Dora Koller, Renato Polimanti

## Abstract

**Background:** Obesity has been associated with more severe clinical manifestations of coronavirus disease 2019 (COVID-19). However, this association can be affected by many correlates of these traits. Due to its large impact on human health, socioeconomic status (SES) could at least partially influence the association between obesity and COVID-19 severity. To estimate the independent effect of traits related to body size and SES on the clinical manifestations of COVID-19, we conducted a Mendelian randomization (MR) study analyzing the effect of obesity-related anthropometric traits on COVID-19 outcomes.

**Methods:** Applying two-sample MR approaches, we evaluated the effects of body mass index (BMI), waist circumference (WC), hip circumference, (HIP) and waist-hip ratio (WHR) studied in up to 234,069 participants from the Genetic Investigation of ANthropometric Traits (GIANT) consortium with respect to three COVID-19 outcomes: severe respiratory COVID-19 (5,101 cases vs. 1,383,241 controls), hospitalized COVID-19 (9,986 cases vs. 1,877,672 controls), and COVID-19 infection (38,984 cases vs. 1,644,784 controls) obtained from the COVID-19 Host Genetics Initiative (HGI). Finally, to test the effect of SES using multivariable MR methods, we analyzed genetic data related to self-reported household income (HI) information from 286,301 UK Biobank (UKB) participants.

**Results:** BMI and WC were associated with severe respiratory COVID-19 (BMI: OR⍰=⍰1.68 p⍰=⍰0.0004; WC: OR⍰=⍰1.72, p⍰=⍰0.007) and COVID-19 hospitalization (BMI: OR=⍰1.62, p⍰=⍰1.35e-06; WC: OR⍰=⍰1.62, p⍰=⍰0.0001). Also, HIP influenced hospitalized COVID-19 (OR⍰=⍰1.31, p⍰=⍰0.012) and COVID-19 infection (OR⍰=⍰1.18, p⍰=⍰0.002). Conversely, HI was associated with lower odds of severe respiratory COVID-19 (OR⍰=⍰0.57, p⍰=⍰0.011) and hospitalized COVID-19 (OR⍰=⍰0.71, p⍰=⍰0.045). Testing these effects in multivariable MR models, we observed that the effect of these obesity-related anthropometric traits on COVID-19 outcomes is not independent of SES effect assessed as HI.

**Conclusions:** Our findings indicate that low SES is a contributor to the observed association between body size and COVID-19 outcomes.

## Background

The coronavirus disease 2019 (COVID-19) resulting from severe acute respiratory syndrome coronavirus 2 (SARS-CoV-2) infection has caused a pandemic since early 2020 [1]. As of May of 2021, this pandemic has led to approximately 168 million COVID-19 cases and more than three million deaths worldwide [2]. The clinical manifestations of COVID-19 vary from asymptomatic infection to a critical illness, i.e., respiratory failure, septic shock, and/or multiple organ dysfunctions [3,4]. Obesity appears to be one of the main factors associated with severe manifestations of COVID-19 [5-8]. Leveraging genetic information, few studies conducted causal inference analyses, showing that BMI and other body measure traits have a putative causal effect on severe and critical COVID-19 illness [9-11]. However, to our knowledge, no investigation was conducted to verify whether the effect of body composition on COVID-19 outcomes was mediated or moderated by other factors. Socioeconomic status (SES) has a large impact on many dimensions of health with individuals with poor SES presenting high morbidity and low life expectancy [12,13]. There is consistent evidence of the impact of SES traits on obesity-related anthropometric traits, such as BMI [14,15]. For example, higher levels of material deprivation have been associated with a higher BMI [16]. Also, higher income has been associated with a higher BMI in developing countries; while in developed countries, BMI has been found to be correlated inversely with median household income [17]. A causal inference analysis based on genetic information showed that BMI may affect social and SES outcomes, but there is also evidence of horizontal pleiotropy (i.e., shared pathways) between these traits [14].

There is a growing literature showing consistently that SES is a primary predictor across COVID-19 outcomes, ranging from infection to mortality [18,19]. Due to its clinical implication, it is important to understand whether the relationship between body size and COVID-19 is affected by their associations with SES. Mendelian randomization studies have demonstrated to be a powerful tool to investigate the potential causative role of SES traits across the spectrum of human health [20-22].

Accordingly, we conducted a MR study to assess the effect of traits related to body size and composition on COVID-19 outcomes, accounting for the effect of SES.

## Methods

### Data Sources

In our study, we investigated the following anthropometric traits: BMI, waist circumference (WC), hip circumference (HIP), and waist-hip ratio (WHR). To assess the effect of body fat distribution, we also investigated BMI-adjusted anthropometric measures, i.e., BMI-adjusted WC, BMI-adjusted HIP, and BMI-adjusted WHR. Genome-wide association statistics were derived from the meta-analyses performed by the Genetic Investigation of ANthropometric Traits (GIANT) consortium. This is an international collaboration aimed to identify genetic loci involved in human body size and shape [23]. These GWAS were conducted in up to 234,069 participants. The cohorts and GWAS procedures are described in previously published articles [24,25].We selected these versions of the GIANT GWAS meta-analyses because they do not include UK Biobank (UKB) cohort, which is a contributor to the COVID-19 data described below and could bias causal inference effect estimates.

We evaluated three COVID-19 outcomes: severe respiratory COVID-19, hospitalized COVID-19, and COVID-19 infection. These data were derived from the release 5 of GWAS meta-analysis conducted by the COVID-19 Host Genetics Initiative (HGI) [26]. This is an international genetics collaboration that aims to uncover the genetic determinants of COVID-19 susceptibility, severity, and outcomes [27]. Analyzed data was obtained only from participants of European descent to population stratification biases. The severe respiratory COVID-19 outcome resulted from the comparison between patients with very severe respiratory failure secondary to COVID-19 (n=5,101) vs. controls (n= 1,383,241). The hospitalized COVID-19 data were generated from the comparison of patients with a laboratory-confirmed SARS-CoV-2 infection that were hospitalized due to COVID-19 symptoms (n=9,986) vs controls (n=1,877,672). Finally, the COVID-19 infection analysis was conducted comparing 38,984 individuals reporting SARS-CoV-2 infection with 1,644,784 controls. Information regarding SARS-CoV-2 infection was derived from a laboratory test, electronic health record, clinically confirmed COVID-19, and self-reported COVID-19 (e.g., by questionnaire) [28].

To investigate the effect of SES, we analyzed genetic data related to self-reported household income (HI) information from UKB participants. HI refers to the combined gross income of all members of a household and was assessed via the touchscreen questionnaire completed by UKB participants [29]. This information was collected using a 5-point scale corresponding to the total household income before tax, 1 being less than £18,000, 2 being £18,000–£29,999, 3 being £30,000–£51,999, 4 being £52,000–£100,000 and 5 being greater than £100,000 [30]. Genome-wide association statistics were obtained from a previous HI GWAS conducted in 286,301 individuals of European descent. GWAS procedure is described in previously published reports [30].

### Mendelian Randomization

MR is an analytic technique to estimate the effect of an exposure on an outcome of interest.10 MR uses genetic variants, which are fixed at conception, to support causal inferences about the effects of risk factors, as they are unlikely to be affected by reverse causation, as they temporally precede the outcome, and confounding factors that act after conception [31]. MR is based on three assumptions: a) the genetic instruments are associated with the outcome of interest; b) the genetic instruments are not associated with potential confounders of the risk factor–outcome association; and c) the genetic instruments affect the outcome only through their effect on the risk factor [32,33].

We estimated the putative causal effect of anthropometric traits on COVID-19 phenotypes using a two-sample MR approach. Leveraging information from genome-wide association statistics, we can estimate the putative causal effect of the exposure on the outcome, which represents the sum of all possible paths from the exposure on the outcome [9,34]. This analysis was conducted using the R package TwoSampleMR [35]. For each anthropometric trait, we defined a genetic instrument based on genome-wide significant variants (p <⍰5⍰×⍰10−8) that were linkage disequilibrium (LD)-independent (r2 <⍰0.001 within a 10,000-kilobase window) based on the 1,000 Genomes Project Phase 3 reference panel for European populations [36]. Also, we excluded those genetic variants that were not present in the COVID-19 outcome GWAS datasets. All analyses were restricted to genome-wide association statistics of European descent to avoid biases due to population stratification.

Our primary MR analysis was conducted using the inverse variance weighted (IVW) approach, because it provides the highest statistical power [37]. To verify the reliability of the IVW results, we compared their concordance with the estimates obtained with the other MR methods: MR-Egger, weighted median, simple mode, and weighted mode [35]. Furthermore, we performed multiple sensitivity analyses (e.g., MR-Egger intercept and heterogeneity tests) described in the Supplementary Methods. To verify the presence of a bidirectional relationship between the anthropometric traits and COVID-19 outcomes, we defined genetic instruments based on genome-wide significant variants (<⍰5⍰×⍰10−8) using the same procedure described above. However, because the number of variants under these criteria was very low (3 SNPs), we also evaluated an alternative approach based on the inclusion of suggestive variants in the instrumental variable and the application of the MR–Robust Adjusted Profile Score (MR-RAPS) approach to account for the possible biases introduced [38]. A detailed description of this approach is available in the Supplementary Methods.

After identifying the putative effects of anthropometric traits, we tested the effect of HI on COVID-19 outcomes, applying the same analytic pipeline. The genetic instrument defined was then used to verify whether the association of anthropometric traits with COVID-19 outcomes is independent of the SES effect. Specifically, we tested the genetic instruments of anthropometric traits and HI with respect to COVID-19 outcomes in a multivariable MR (MVMR) analysis. This is an extension of the standard MR framework to consider multiple potential risk factors in a single model and calculate the independent association of each risk exposure with the outcome [33,39,40]. The MVMR analysis was conducted using the MendelianRandomization R package [39].

## Results

We observed consistent associations of anthropometric traits on COVID-19 outcomes (Figure 1). The strongest association was observed with respect to BMI and WC: genetically determined BMI, and WC was associated with increased odds of severe respiratory COVID-19 (BMI: OR=1.68, p=0.0004; WC: OR=1.72, p=0.0072) and hospitalized COVID-19 (BMI: OR=1.62, p=1.35e-06; WC: OR=1.62, p=0.0001). Reduced effects were observed for these anthropometric traits with respect to COVID-19 infection (BMI: OR=1.26, p=1.16e-05; WC: OR=1.23, p=0.001). Conversely, genetically determined HIP showed a positive association with hospitalized COVID-19 (OR=1.31, p⍰=0.012) and COVID-19 infection (OR=1.18, p=0.0016), but not with severe respiratory COVID-19 (OR=1.24, p=0.186). The IVW effect estimate was consistent with the ones estimated with all other MR methods estimates and no heterogeneity or horizontal pleiotropy was observed within BMI, WC, and HIP genetic instruments (Supplemental Table 1). Also, we found a positive association between WHR and COVID-19 infection (OR=1.39, p=0.005) only with the weighted median method. However, we detected the presence of two potential outliers within the genetic instrument in the leave-one-out test in the association WHR and COVID-19 infection (rs9937053 and rs998584) (Supplemental Table 2). After removal of the variants contributing to heterogeneity, the association of genetically determined WHR with COVID-19 infection was not significant (OR=1.08, p=0.361) with any of the evaluated MR methods. Among the anthropometric traits related to body fat distribution (i.e., WHR and traits adjusted for BMI), genetically-determined BMI-adjusted HIP was positively associated with COVID-19 infection (OR=1.11, p=0.0129).

**Figure 1.**
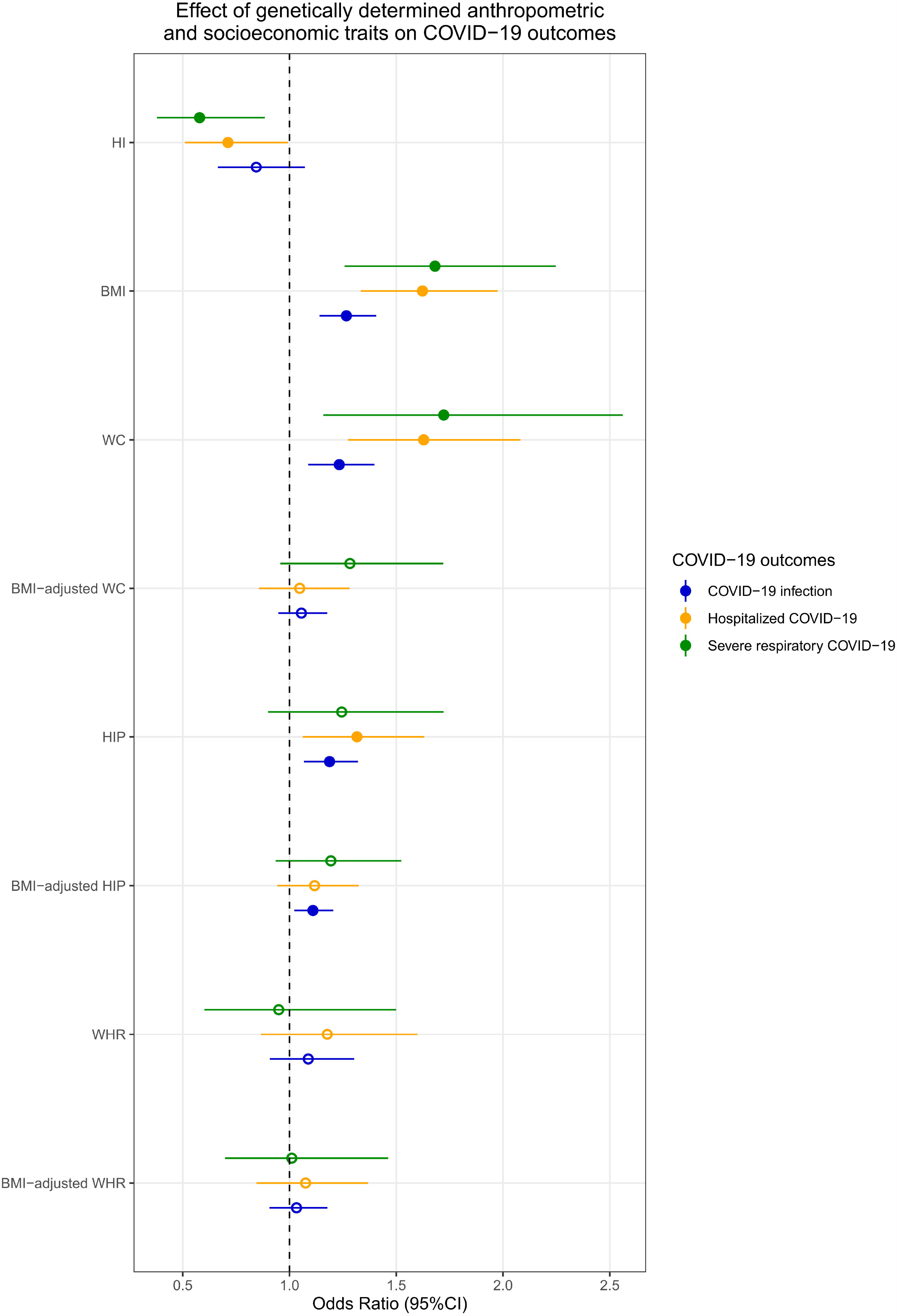
Results of the MR analysis testing the effect of genetically determined anthropometric and socioeconomic traits on severe respiratory COVID-19 (green), COVID-19 hospitalization (orange), COVID-19 infection (blue). Effect size and 95% confidence interval are reported for each MR test. Significant associations are denoted by filled shapes. Abbreviations: body mass index (BMI); waist circumference (WC); hip circumference (HIP); waist-hip ratio (WHR); Confidence Interval (CI).

To verify whether the effects of BMI, WC, and HIP on COVID-19 outcomes were independent of each other, we entered the genetic instruments in a MVMR model. The effects observed in the univariate MR were null after accounting for the effect of other anthropometric traits (Supplemental Table 3).

We also tested the possible reverse association of COVID-19 outcomes on the anthropometric traits that showed significant effects in the initial MR analyses (Figure 1). The analysis based on genetic instruments considering genome-wide significant variants showed null effects (p>0.05). In the analysis based on suggestive variants, we observed an effect of genetically-determined hospitalized COVID-19 on BMI (β=0.01, p=0.0062). However, the effect size is much smaller than the one observed in the reverse direction (β_Hospitalized COVID-19→BMI_=0.01 vs. β_BMI→Hospitalized COVID-19_=0.484). Additionally, we found evidence of strong heterogeneity within the genetic instruments evaluated (Supplemental Table 4). Since the leave-one-out analysis did not indicate the presence of major outliers, we hypothesized that the heterogeneity observed could be the result of the widespread horizontal pleiotropy among the variants included in the genetic instruments. Thus, we re-estimated the effects using the MR-RAPS approach with the squared error loss method (I2) to account for overdispersion (systematic pleiotropy) within the genetic instrument [37]. The result observed was consistent with the IVW estimates (β=0.01, p=0.013) (Supplemental Figure 1). Complete results from the reverse MR analysis are available in Supplemental Table 4 (Supplementary Material 1).

To test whether the effect of anthropometric traits on COVID-19 is independent of SES, we initially verified the reliability of the HI genetic instrument with the univariate MR approach. There was a protective effect of HI on severe respiratory COVID-19 (OR=0.58, p=0.011) and hospitalized COVID-19 (OR=0.71, p=0.045). These estimates were not affected by heterogeneity or horizontal pleiotropy (Supplemental Table 5). Because of the UKB cohort was included in both HI and COVID-19 GWAS, we verified that this sample overlap did not affect the estimates observed. Using COVID-19 HGI meta-analysis excluding UKB sample, we observed no statistical difference in the effect size of the associations observed with respect to the analyses obtained using the COVID-19 HGI meta-analyses including UKB sample (Supplemental Table 6). Similar to BMI, the effect of severe respiratory COVID-19 on HI was significant (β=0.006, p=4.29e-5) but much smaller and with opposite sign than the one observed in the initial MR test (β_Severe respiratory COVID-19→HI_=0.006 vs. β_HI→ Severe respiratory COVID-19_=⍰-0.546) (Supplemental Table 7). In the MVMR, the effect of the anthropometric traits on severe respiratory COVID-19 (i.e., BMI and WC) and hospitalized COVID-19 (i.e., BMI, WC, and HIP) was null when accounting for the effect of HI (Figure 2; Supplemental Tables 8 and 9). Conversely, the effect of HI on severe respiratory COVID-19 and hospitalized COVID-19 was still significant when accounting for each anthropometric trait individually and including all anthropometric traits in the same MVMR model (Supplemental Tables 8, 9, and 10).

**Figure 2.**
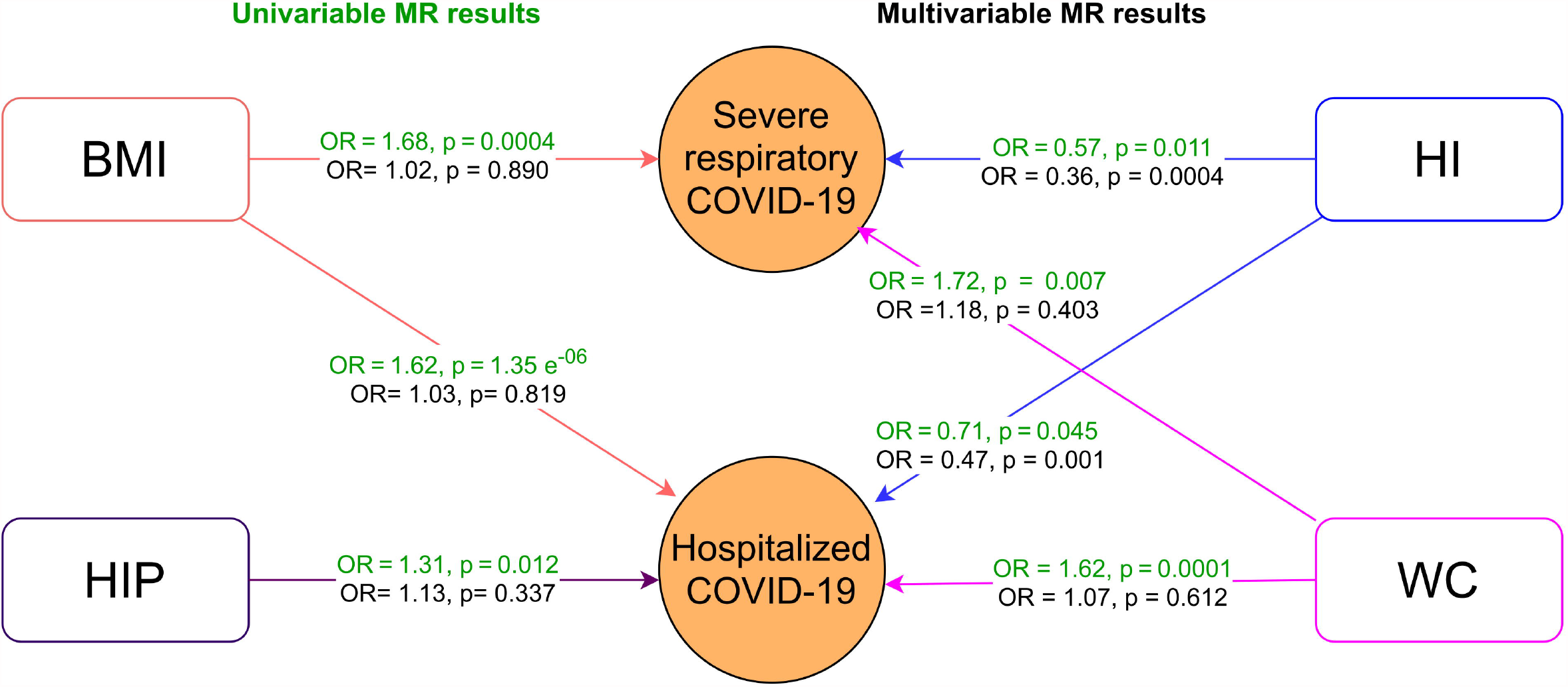
Multivariable Mendelian randomization. Arrows indicate the direction of the effect between the traits. ORs and p-values from each association are shown in the middle of the arrows for the univariate MR analysis (in green) and the multivariable MR analysis (in black). For the anthropometric traits, we report the estimates related to the MVMR model with household income (HI; Supplemental Tables 8 and 9). For HI, we report the estimated related to the MVMR model including all anthropometric traits with significant effect in the univariable MR analysis (Supplemental Table 10). COVID-19 phenotypes are denoted by circles, while anthropometric and socioeconomic traits are denoted by rectangles. Abbreviations: household income (HI); body mass index (BMI); waist circumference (WC); hip circumference (HIP); Odds ratio (OR); p-value (p).

## Discussion

The association of obesity-related traits with COVID-19 outcomes has been confirmed by multiple investigations, also including MR analyses that showed putative causal effects of BMI [9,10,11,41,42], WC [9,10], and trunk fat ratio [9]. Some of these previous investigations also have included MVMR analyses that highlighted how the effect of these traits on COVID-19 is independent of several cardiometabolic traits and other known risk factors [43]. However, to our knowledge, no study evaluated whether the effect of obesity-related traits on COVID-19 outcomes is independent of SES. Because of the unprecedented impact of COVID-19 on individuals and society, it is crucial to identify individuals at risk and relevant modifiable factors to reduce COVID-19 morbidity and mortality. Current findings support that traits related to body size should be among the primary targets to prevent COVID-19 severe symptoms. However, there is a well-established relationship between body size variation and SES that may affect the associations observed [44].

The results of our univariable MR analysis showed positive associations between genetically determined BMI with COVID-19 outcomes. These findings are consistent with previous reports of a causal impact of BMI on severe respiratory and hospitalized COVID-19 [9,10,11,41,42]. However, our multivariable analysis indicated that these associations are mediated by the effect of SES on COVID-19 outcomes. These findings have major public health implications, because while previous studies highlighted the importance to consider obesity among the risk factors associated with COVID-19 severe outcomes, our results strongly highlight that this evidence should be put in the context of the effect of SES on both body size and COVID-19. The mechanisms by which HI is mediating the association between obesity-related anthropometric traits and COVID-19 might be similar to those proposed for other known health outcomes. In general, low-income populations have reduced access to medical care, with subsequent worse health at baseline and lower opportunity to receive adequate treatment to health complications in comparison with high-income groups [45,46]. Finally, these healthcare inequalities are translated into a higher morbidity and mortality risk in communities with a lower household income [47].

Regarding anthropometric traits, individuals with a lower SES generally have a higher BMI and increased risk of obesity [15]. There are multiple social and environmental factors contributing to the association between SES and BMI, i.e., economic difficulties, the affordability of energy-dense foods, low dietary quality, poor health literacy, occupational status, lifestyle behaviors, such as low physical activity and sedentary behaviors [48-53]. Furthermore, factors influencing both SES and obesity that also act as modulators of their association have been identified, such as sex, ethnicity, country’s income economy, and education [54,55]. For example, in developed countries, obesity is more prevalent in individuals with lower SES, while in developing countries there is not a clear trend [54,55]. Also, generally for men the obesity prevalence similar at all income levels. However, non-Hispanic African-American and Mexican-American males with higher income are more likely to be obese compared with those with low income. Conversely, high-income women are less likely to be obese than low-income women [56]. Despite the relevance of the mentioned environmental and social factors, it is important to consider that obesity is a complex multifactorial disease influenced by both environmental and genetic factors, as well as by the interaction between them [48].

Similar to obesity, besides environmental factors there is evidence of the effect of genetic factors on SES, particularly on HI [30,57-59]. The genetic variants associated with a higher income were functionally linked with GABAergic and serotonergic neurotransmission and linked to cognitive abilities [30].

In our study, we found that genetically determined HI had a negative causal effect on both very severe respiratory confirmed COVID-19 and hospitalized COVID-19. Previously, a longitudinal study reported a negative relationship between HI and COVID-19 mortality, which might be considered an extreme COVID-19 phenotype [45]. Interestingly, there was no difference in COVID-19 infection rate among the different income groups [45]. Hence, these previous results and ours suggest that HI influences only COVID-19 severe phenotypes. Further exploration of the genetic variants involved in this association might provide information on their biological role in the predisposition to COVID-19 complications. The elucidation of the mechanisms will contribute to the design of strategies and/or treatments that could modulate their effect on COVID-19 outcomes. The mechanisms linking the genetic predisposition to obesity and COVID-19 are likely to be multifactorial and warrant clarification in further research [59].

Overall, the anthropometric traits evaluated here showed a consistent effect on COVID-19 phenotypes. However, this association is not direct and multiple factors might be acting as confounders or mediators in these associations i.e., health behaviors associated with SES such as physical inactivity, diet, tobacco, and alcohol use [60]. Although the results of the reverse MR might suggest an effect of genetically predicted COVID-19 phenotypes on anthropometric traits and SES, the effect size is negligible, suggesting a possible residual effect of horizontal pleiotropy. The identification of factors underlying these causal associations will contribute to the design of adequate risk-stratification instruments for patients with COVID-19. Furthermore, it might contribute on a higher scale to the design of public policies oriented to the prevention of adverse outcomes in vulnerable groups. Although these public policies will not modify the genetic factors involved in the development of severe COVID-19 symptoms, they might impact modifiable risk factors and reduce social inequalities. For example, strategies that improve the access to adequate medical services might lead to the adverse impact of COVID-19 in low-income communities [45].

The limitations of the present study should be acknowledged. First, this study was conducted using data generated from participants of European descent. Accordingly, the results obtained may not be generalization to populations with other ancestral origins. Further research is necessary to evaluate the effect of anthropometric and socioeconomic traits in populations with diverse ancestral background. Second, in this study we could not evaluate the presence of sex-specific effects as large-scale datasets informative of sex-specific COVID-19 susceptibility are not available at this time. There are known sex differences in the mechanisms underlying the association of SES with body size and composition [14,61]. Therefore, future sex-stratified studies are required to understand the processes linking these traits with COVID-19 outcomes. Third, although we conducted multiple sensitivity analyses, we cannot discard completely the influence of potential confounders in our results. Therefore, complementary studies are needed to confirm and further explore the findings here reported.

## Conclusions

We provide the first evidence that the relationship of obesity-related anthropometric traits with COVID-19 outcomes is not independent of SES. This result has major public health implications because it supports that preventive strategies targeting body size and composition to reduce COVID-19 morbidity and mortality may not be effective if they are not considered in the context of SES.

## Data Availability

All data discussed in this study are provided in the article and in the Supplementary Material.

## List of abbreviations

BMI: body mass index
WC: waist circumference
HIP: hip circumference
WHR: waist-hip ratio
COVID-19: coronavirus disease 2019
SES: socioeconomic status
MR: Mendelian randomization
GIANT: Genetic Investigation of ANthropometric Traits
UKB: UK Biobank
SARS-CoV-2: severe acute respiratory syndrome coronavirus 2
HI: household income
HGI: COVID-19 Host Genetics Initiative
GWAS: genome-wide association studies
LD: linkage disequilibrium
OR: odds ratio

## Declarations

### Ethics approval and consent to participate

This study was conducted using genome-wide association statistics generated by previous studies. Owing to the use of previously collected, deidentified, aggregated data, this study did not require institutional review board approval.

### Consent for publication

Not applicable

### Availability of data and materials

The datasets used in this study are available in the GIANT consortium website (http://portals.broadinstitute.org/collaboration/giant/index.php/GIANT_consortium), the COVID-19 Host Genetics Initiative website (https://www.covid19hg.org/) and the UK Biobank website (https://www.ukbiobank.ac.uk/). All data discussed in this study are provided in the article and in the Supplementary Material.

## Authors’ contributions

BCM and RP participated in the concept and design of the study, statistical analysis, and drafting of the manuscript. BCM, FW, GP, FDA, ADL, DK, RP participated in the acquisition, analysis, interpretation of data, and critical revision of the manuscript for important intellectual content. FW, DK, RP obtained the funding for this study. FW, GP, FDA, ADL, DK, RP provided the administrative, technical, and material support for this study. RP supervised the present study. All the authors revised and approved the final version of this manuscript.

## Acknowledgements

We thank study participants and research groups contributing to the GIANT Consortium and the COVID-19 HGI for making their data available.

## Supplementary information

### Supplementary Material 1

Supplementary Material 1. doc

Supplemental Figure 1. Results of the MR analysis testing the effect of genetically determined severe respiratory COVID-19, hospitalized COVID-19, reported COVID-19 infection on socioeconomic and anthropometric traits.

### Supplementary Methods

Supplementary. Methods.docx

1. MR Sensitivity analyses
2. Alternative approach to select genetic instruments in the MR analysis to test the effect of COVID-19 outcomes in anthropometric traits

### Supplementary Tables

Supplementary.Tables.xlsx

Supplementary Table 1. Effect of anthropometric traits on COVID-19 outcomes

Supplementary Table 2. Supplemental Table 2. Leave-one out results from the MR analysis evaluating the effect of WHR on COVID-19.

Supplementary Table 3. Multivariable MR results for the effect of anthropometric traits on COVID-19 outcomes

Supplementary Table 4. Effect of COVID-19 Phenotypes on anthropometric traits

Supplementary Table 5. Effect of socioeconomic status on COVID-19 Phenotypes

Supplementary Table 6. Difference between IVW estimates for HI with respect to COVID-19 HGI GWAS meta-analysis with and without UK Biobank

Supplementary Table 7. Effect of COVID-19 Phenotypes on socioeconomic status

Supplementary Table 8. Multivariable MR results for the effect of anthropometric traits and socioeconomic status on severe respiratory COVID-19

Supplementary Table 9. Multivariable MR results for the effect of anthropometric traits and socioeconomic status on hospitalized COVID-19

Supplementary Table 10. Multivariable MR results for anthropometric traits and socioeconomic status on COVID-19 outcomes

