## Supplementary Figure 1 for "The effect of obesity-related traits on COVID-19 severe respiratory symptoms is mediated by socioeconomic status: a multivariable Mendelian randomization study"

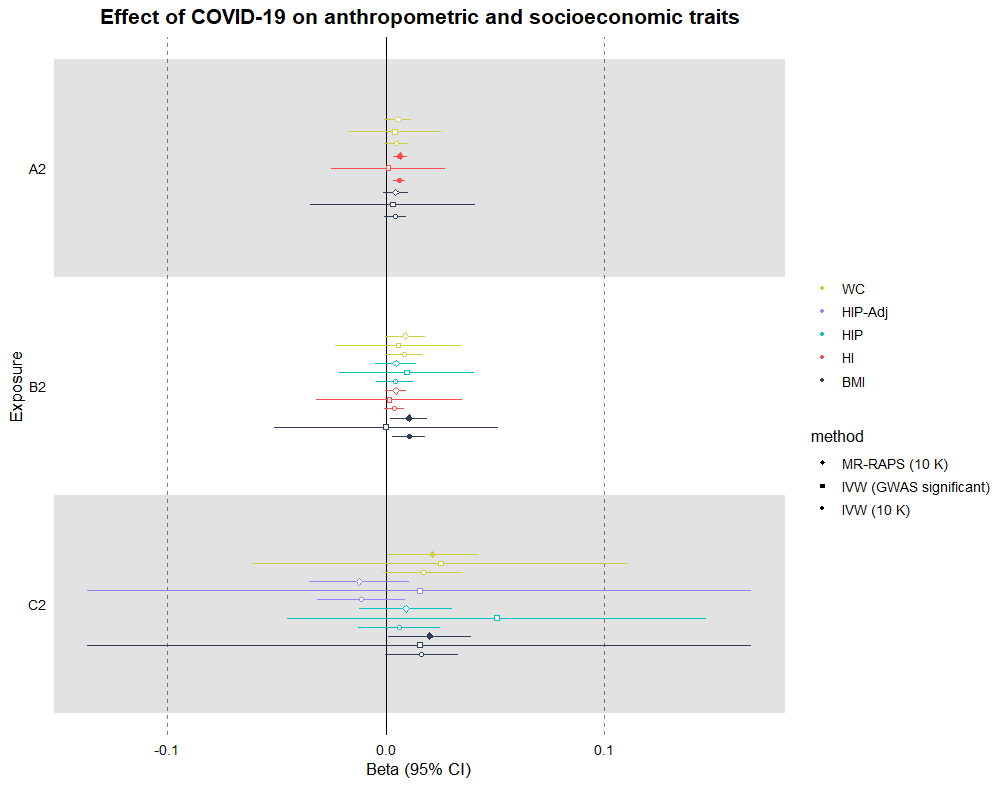

**Supplemental Figure 1.** Results of the MR analysis testing the effect of genetically determined severe respiratory COVID-19 (A2), hospitalized COVID-19 (B2), reported COVID-19 infection (C2) on socioeconomic and anthropometric traits. Effect size (beta) and 95% confidence interval are reported for each MR test. Significant associations are denoted by filled shapes. Abbreviations: severe respiratory COVID-19 (A2), hospitalized COVID-19 (B2), reported COVID-19 infection (C2); household income (HI); Townsend Social deprivation Index (TO); body mass index (BMI); waist circumference (WC); hip circumference (HIP); waist-hip ratio (WHR); body mass index -adjusted waist circumference (WC-Adj); body mass index -adjusted hip circumference (HIP-Adj); body mass index -adjusted waist-hip ratio (WHR-Adj); Confidence Interval (CI); inverse variance weighted (IVW).
