## Supplementary Methods for "The effect of obesity-related traits on COVID-19 severe respiratory symptoms is mediated by socioeconomic status: a multivariable Mendelian randomization study"

**MR Sensitivity analyses**

Additionally, the presence of horizontal pleiotropy and heterogeneity within the genetic instruments, which are potential confounders in MR analyses, was tested using MR-Egger regression intercept (1), and IVW and MR-Egger heterogeneity tests (2), respectively. Furthermore, outliers contributing to the heterogeneity (variability in the causal estimates obtained for each SNP), and horizontal pleiotropy (the variant affects the disease indirectly, i.e., outside its effect on the exposure) within the genetic instruments were identified by leave-one-out analyses and the visual assessment of forest and funnel plots. The genetic variants identified as outliers were removed from the analysis and the causal estimates were recalculated to evaluate the potential biases generated by these outliers in the MR results. Similar to other MR studies (3-5) we avoided inference based simply on p value thresholds. The direction and strength of effect for each MR association, together with the corresponding p value, was considered to better reflect the spectrum of evidence related to these results (6).

**Alternative approach to select genetic instruments in the MR analysis to test the effect of COVID-19 outcomes in anthropometric traits**

Within COVID-19 HGI release-5 data, the largest sample is available for the meta-analysis including data from 23andMe in addition to the other cohorts included in the analyses already described in the Data Source section. In this large dataset, COVID-19 was investigated in 5,101 cases and 1,383,241 controls for the A2 phenotype; 9,986 cases and 1,877,672 controls for B2 phenotype; and 38,984 cases and 1,644,784 controls for C2 phenotype. However, due to 23andme data sharing restrictions, genetic association statistics are available only for the top 10,000 variants. Accordingly, we extracted the LD-independent loci among them and used this information to define genetic instruments for the COVID-19 outcomes investigated. These genetic instruments including variants not reaching genome-wide significance might lead to the violation of the MR assumptions. Thus, we considered as primary analysis the estimates obtained from the IVW method and the MR–Robust Adjusted Profile Score (MR-RAPS) approach. The MR-RAPS approach estimates the causal effect under pervasive horizontal pleiotropy and is robust to occasional outliers (7). The reliability of the findings was tested using multiple sensitivity analyses described above.
